# The Impact of Endoscopic Ultrasound Adoption on Etiological Shifts in Biliary Obstruction: A 15-Year Real-World Study

**DOI:** 10.64898/2026.05.06.26352511

**Authors:** Ningyuan Wen, Nianheng Wu, Haojun Wu, Haili Zhang, Yufu Peng, Hongwei Xu, Yonggang Wei

## Abstract

**Background and Objectives:** The etiology of biliary obstruction has undergone notable shifts over recent decades, yet long-term epidemiological studies addressing these changes remain scarce. With the widespread clinical adoption of endoscopic ultrasound (EUS), its role in altering diagnostic patterns warrants investigation. This study aimed to characterize the evolving disease patterns of biliary obstruction and specifically evaluate the impact of EUS adoption on driving these perceived etiological shifts over a 15-year period.

**Methods:** This retrospective, single-center study analyzed data from patients with biliary obstruction over a 15-year period. Time-series analysis was employed to characterize evolving disease patterns. To investigate the drivers underlying the observed trends, we applied a difference-in-differences (DID) analytical framework, uniquely treating the widespread clinical adoption of EUS as a natural experiment. Furthermore, multivariable logistic regression was utilized to identify independent predictors for malignant biliary obstruction of pancreatic origin.

**Results:** Among 5,672 patients with pathological diagnoses, the disease spectrum shifted significantly toward malignant etiologies, particularly pancreatic and ampullary cancers, over the study period. The DID analysis confirmed that the broad adoption of EUS was associated with a significant relative increase in the precise diagnosis of malignancies detectable by this modality. Multivariable analysis further identified the EUS promotion era and calendar year as independent predictors for the pancreatic origin of malignancy.

**Conclusions:** The observed increase in pancreatic and ampullary cancers among patients with biliary obstruction is significantly associated with the enhanced diagnostic capabilities brought by EUS. This suggests that the diagnostic evolution driven by the widespread adoption of EUS, alongside potential epidemiological changes, is a major contributing factor to the perceived etiological shifts in biliary obstruction.

## Introduction

Biliary obstruction is a common clinical syndrome with significant morbidity and mortality, often presenting with jaundice, cholangitis, or progressive liver dysfunction^[1]^. The etiological spectrum is broad, encompassing benign causes such as choledocholithiasis and benign strictures, as well as highly lethal malignancies including pancreatic cancer and cholangiocarcinoma^[2-5]^. Accurate and timely etiological diagnosis is paramount, as it critically influences patient prognosis and dictates subsequent therapeutic pathways, which vary dramatically between benign and malignant conditions^[6]^.

Despite its clinical importance, long-term temporal trends in the definitive etiology of biliary obstruction remain poorly characterized. Existing knowledge is often fragmented, relying on smaller cohorts or shorter observation periods, leaving a significant gap in our understanding of the true longitudinal evolution of this disease spectrum^[7,8]^. While a clinical perception exists that the proportion of malignant causes is increasing, it has been uncertain whether this reflects a true epidemiological shift or is a diagnostic artifact^[9]^. Critically, the past two decades have not only seen advances in imaging but have heralded a paradigm shift in our ability to obtain definitive pathological confirmation. This evolution has been driven by the widespread adoption and technical refinement of advanced endoscopic procedures for tissue acquisition. While Endoscopic Retrograde Cholangiopancreatography (ERCP) has long provided a conduit for brush cytology and intraductal biopsy, the ascent of Endoscopic Ultrasound-guided Fine-Needle Aspiration and Biopsy (EUS-FNA/B) has been particularly transformative, offering a highly sensitive and safe modality to histologically diagnose lesions, especially solid pancreatic masses, that were previously challenging to sample^[10,11]^. This enhanced capability for pathological diagnosis, rather than just imaging suspicion, fundamentally complicates the interpretation of observed temporal trends and creates a critical need to disentangle the impact of technology from true changes in disease patterns.

To address this knowledge gap, we began by conducting a large-scale, retrospective time-series analysis of all patients presenting with biliary obstruction at our tertiary center over a continuous 15-year period. The primary aim of this initial analysis was to systematically characterize the temporal trends in the etiological spectrum and diagnostic patterns. Our findings revealed a significant shift in the disease spectrum, most notably a rising proportion of diagnosed malignant obstructions, particularly those of pancreatic origin. This observation prompted the central question of our study: what is the underlying driver of this shift? We therefore proceeded to investigate the impact of technological advancement, hypothesizing that the widespread adoption of EUS was a key contributor. By leveraging this EUS adoption period as a natural experiment within a Difference-in-Differences (DID) framework, we sought to quantify its specific impact and ultimately answer the question: Has the disease changed, or have we simply learned to see it better?

## Patients and Methods

### Participants

This single-center, retrospective cohort study was conducted at West China Hospital of Sichuan University, a major tertiary referral center in western China. The study protocol was approved by the Ethics Committee on Biomedical Research, West China Hospital of Sichuan University, and the requirement for individual informed consent was waived due to the retrospective nature of the analysis. The study was preregistered in the Open Science Framework (registration DOI: https://doi.org/10.17605/OSF.IO/JYDS3 ) to ensure transparency.

We identified all hospitalized patients with a clinical diagnosis of biliary obstruction between January 1, 2008, and January 31, 2023, by searching the hospital’s electronic health record (EHR) database. The initial search was based on International Classification of Diseases, 10th Revision (ICD-10) codes related to biliary obstruction, jaundice, and diseases of the biliary tract and pancreas, as well as keyword searches in discharge summaries. The final cohort for analysis was established based on the following criteria:

Inclusion Criteria: (1) hospitalized patients with a discharge diagnosis of biliary obstruction during the study period; (2) objective evidence of biliary obstruction, defined as ductal dilation or a clear obstructive lesion confirmed by at least one of the following imaging modalities: abdominal ultrasound, contrast-enhanced computed tomography (CECT), magnetic resonance imaging/magnetic resonance cholangiopancreatography (MRI/MRCP), or endoscopic ultrasound (EUS).

Exclusion Criteria: (1) Patients with incomplete or unavailable critical data, including definitive imaging reports or final etiological diagnosis.

A detailed flowchart illustrating the patient selection process is provided in Figure 1.

**Figure. 1.**
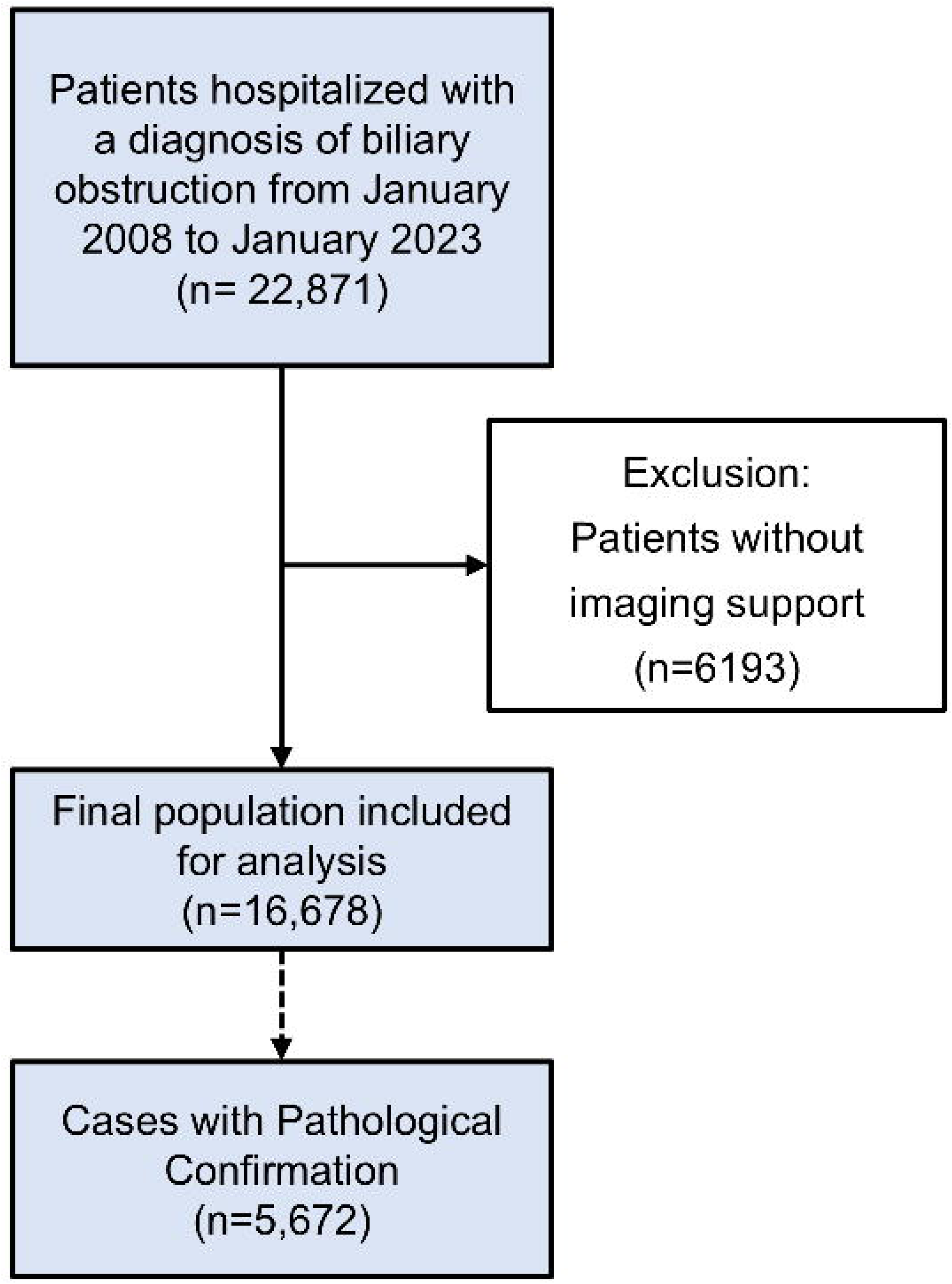
Flowchart of study population selection.

### Collection of clinical data

A dedicated team of trained researchers extracted and curated data from the institution’s electronic health record (EHR) database using a standardized data collection form. To ensure data quality and consistency, a subset of records was independently reviewed by two investigators to resolve any discrepancies. All patient data were de-identified prior to analysis. The collected variables included baseline demographics (age at admission, sex) and key laboratory results upon presentation. Documented details of the pathological confirmation process were extracted, including the specific method used to obtain tissue, such as endoscopic ultrasound-guided fine-needle biopsy (EUS-FNB), endoscopic retrograde cholangiopancreatography (ERCP) with tissue sampling, percutaneous biopsy, or surgical resection, along with the date each definitive procedure was performed. The primary outcome was the final etiology of biliary obstruction, which was confirmed either by the gold standard of pathological examination.

### Statistical analysis

All statistical analyses were performed using R software (version 4.4.2), with a two-sided P-value < 0.05 considered statistically significant. Baseline characteristics were summarized using standard descriptive statistics. To analyze temporal changes over the 15-year study period, we visualized trends in etiologies and imaging utilization with time-series plots fitted with a Locally Estimated Scatterplot Smoothing (LOESS) algorithm and quantitatively assessed for monotonicity using Spearman’s rank correlation. A multivariable logistic regression model, developed with a backward stepwise selection method based on the Akaike Information Criterion (AIC), was used to identify independent predictors of a malignant etiology. Furthermore, a Difference-in-Differences (DID) quasi-experimental analysis was employed to specifically evaluate the impact of the widespread adoption of advanced imaging technologies on diagnostic outcomes, comparing changes over time between malignant and benign cohorts to isolate the technology’s effect from other temporal trends^[12]^.

## Results

### Baseline characteristics of the study population

During the study period from 2008 to 2023, a total of 16,678 patients were admitted for biliary obstruction. Of these, 5,672 (34.0%) had a definitive pathological confirmation and were included in the final analysis. The median age of this cohort was 58.4 years, and the population was predominantly male (56.8%). Both age and sex distribution remained stable across the two study periods (2008–2015 vs. 2016–2023; p=0.24 and p=0.88, respectively). A comprehensive summary of patient characteristics is provided in Table 1.

Notably, the number of patients with pathologically confirmed diagnoses increased substantially, from 1,262 in the 2008–2015 period to 4,410 in the 2016–2023 period. Overall, malignant diseases (n=3,683, 64.9%) were more common than benign conditions (n=1,978, 34.9%). A significant temporal shift was observed in the disease spectrum, with the proportion of malignant cases increasing from 58.6% in the earlier period to 66.7% in the later period (p<0.001). This was accompanied by a changing etiological landscape (p<0.001). Specifically, the proportion of common bile duct stones decreased (from 25.0% to 16.4%), while the proportions of pancreatic ductal adenocarcinoma (from 14.9% to 17.8%) and hilar cholangiocarcinoma (from 7.1% to 10.8%) increased over time.

Furthermore, the utilization of Endoscopic Ultrasound (EUS) for diagnostic purposes saw a significant increase, rising from 11.0% to 16.3% (p<0.001). Patients in the later period also presented with significantly higher median levels of total bilirubin (68.0 vs. 53.7 μmol/L), direct bilirubin (41.3 vs. 32.0 μmol/L), and CA 19-9 (82.6 vs. 45.3 U/mL) compared to those in the earlier period (all p<0.001).

### Temporal trends in pathological confirmation rates and etiological spectrum of biliary obstruction

Over the 16-year study period, we observed a significant and steady increase in the proportion of patients with biliary obstruction who received a definitive pathological diagnosis (Figure 2). The absolute number of patients with a pathological diagnosis increased more than tenfold, from 108 in 2008 to 1,699 in 2023. Consequently, the annual proportion of patients undergoing pathological confirmation rose from 30.6% in 2008 to 43.8% in 2023 (Figure 2A). This clear upward trend was statistically significant, as demonstrated by a strong positive correlation between the year and the pathological diagnosis rate (Pearson’s r = 0.69, p = 0.003; Figure 2B), underscoring a progressively improving diagnostic capability over time.

**Figure. 2.**
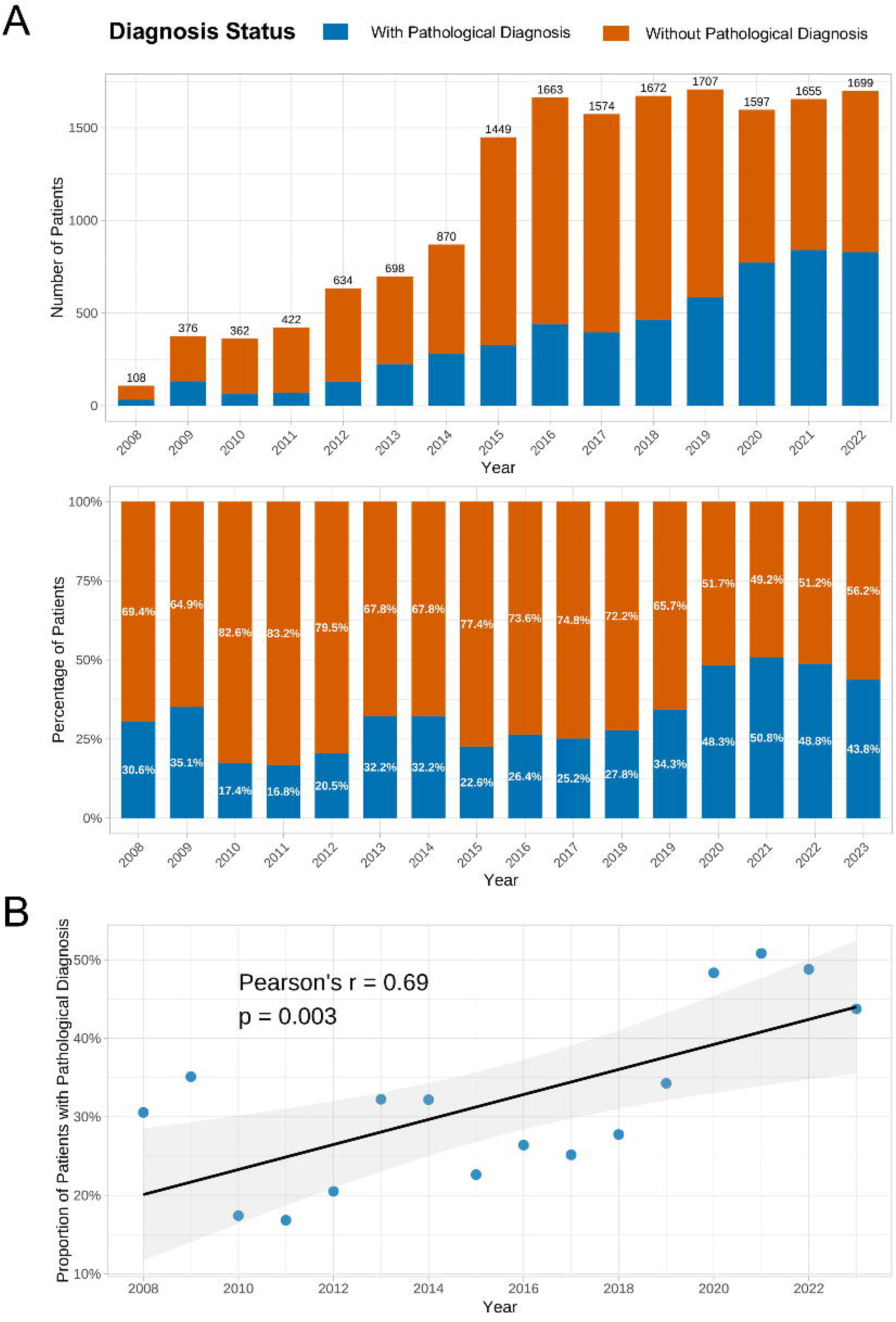
Trends in the application of pathological diagnosis for biliary obstruction from 2008 to 2023. **(A)** Annual absolute numbers (top panel) and percentage composition (bottom panel) of patients with and without a pathological diagnosis. The numbers above the bars in the top panel indicate the total number of patients with biliary obstruction each year. **(B)** Correlation analysis between the year and the proportion of patients receiving a pathological diagnosis. The plot shows the linear regression line with its 95% confidence interval, along with the Pearson’s correlation coefficient (r) and the corresponding p-value.

Coinciding with this improved diagnostic confirmation, the etiological spectrum of biliary obstruction among pathologically confirmed cases underwent a significant transformation (Figure 3). The most prominent trend was the shift towards a higher proportion of malignant etiologies (Figure 3C). The proportion of malignant cases demonstrated a strong and statistically significant linear increase over the study period, rising from 57.6% in 2008 to 71.4% in 2023 (Adj. R² = 0.842, p < 0.001; Figure 3D). This overall shift was driven by changes in the composition of specific diseases (Figure 3A). Notably, the proportional contribution of pancreatic cancer and hilar cholangiocarcinoma increased over time. Conversely, the proportion of benign causes, primarily represented by biliary stones, showed a marked decline. This evolving etiological landscape was also reflected in the anatomical site of obstruction, with a growing proportion of blockages located in the pancreas and perihilar regions over the years (Figure 3B).

**Figure. 3.**
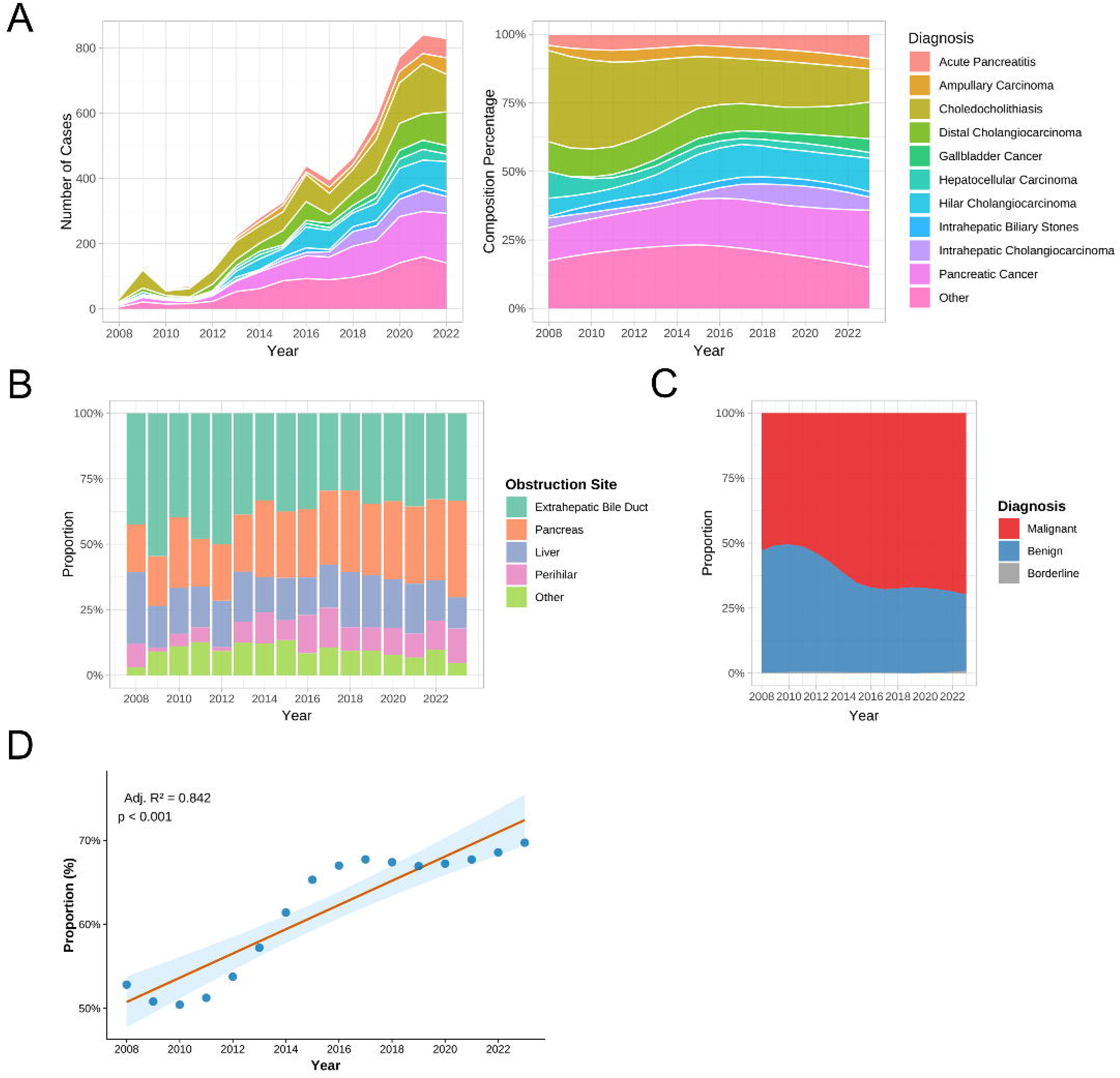
Etiological composition and trends of malignant diagnoses in patients with biliary obstruction. **(A)** Annual absolute numbers (left panel) and composition percentage (right panel) of the final diagnoses. **(B)** Temporal trends in the proportional composition of different obstruction sites. **(C)** Temporal trends in the proportional composition of malignant, borderline, and benign diagnoses. **(D)** Linear regression analysis showing the increasing proportion of malignant diagnoses over the years. The adjusted R-squared (Adj. R²) and p-value are displayed.

### Impact of EUS adoption on diagnostic patterns

#### Temporal trends in the adoption of EUS-guided biopsy

To understand the drivers behind the increasing rate of pathological confirmation, we analyzed the temporal trends of different diagnostic methods (Figure 4). While surgical resection remained the most common method for obtaining a pathological diagnosis throughout the study period, its proportional contribution decreased from 78.8% in 2008 to 63.1% in 2023. In contrast, the use of EUS-guided biopsy experienced a dramatic expansion.

**Figure. 4.**
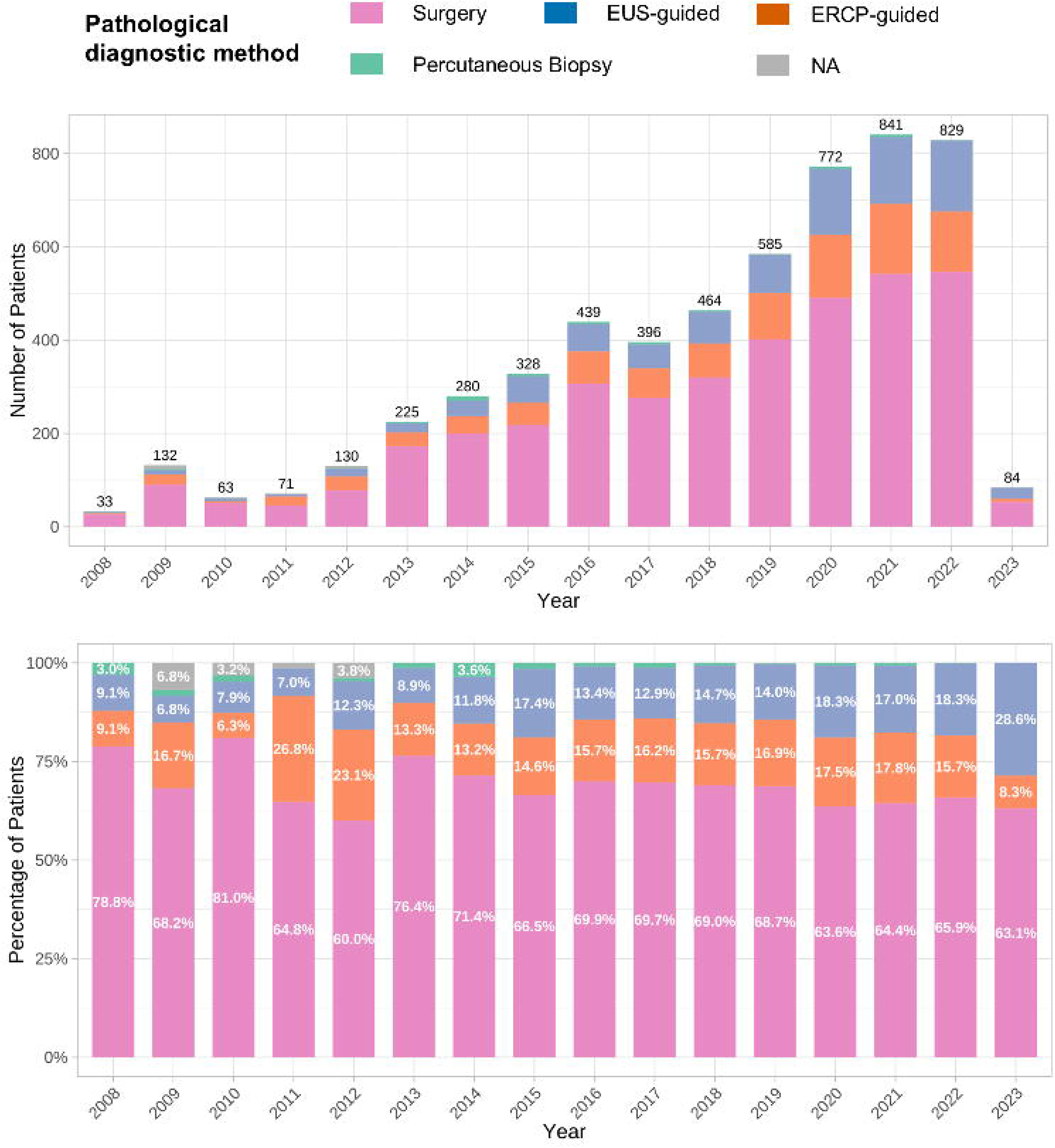
Application trends of different pathological diagnostic methods from 2008 to 2023. The figure illustrates the annual absolute numbers (top panel) and composition percentages (bottom panel) of patients who obtained a pathological diagnosis via different methods, including surgery, EUS-guided procedures, ERCP-guided procedures, and percutaneous biopsy.

As shown in Figure 4, the absolute number of EUS-guided procedures was negligible in the early years but began a rapid ascent around 2015. This translated into a significant shift in its proportional use, rising from just 9.1% in 2008 to a peak of 28.6% of all pathologically confirmed cases in 2023. This growth trajectory of EUS aligns closely with the previously observed increase in the overall pathological confirmation rate.

#### Impact of widespread EUS adoption on the diagnostic spectrum: a Difference-in-Differences (DID) analysis

Given the strong temporal correlation between the rise of EUS and the shift in the etiological spectrum, we employed a Difference-in-Differences (DID) analysis to formally test for a causal link. We categorized malignancies based on their anatomical location and accessibility to EUS biopsy. Pancreatic and ampullary cancers were defined as the “EUS-sensitive” group, while perihilar and intrahepatic leisions, which are less amenable to standard EUS-guided biopsy, were defined as the “EUS-insensitive” group. We set 2015 as the intervention point, marking the beginning of the “EUS Promotion Era” when the technology became widely adopted in our institution.

Visual inspection of the trends prior to 2015 supported the parallel trends assumption, with both groups showing relatively stable and parallel trajectories (Figure 5A). Following the intervention point, a striking divergence occurred. The proportion of diagnosed EUS-sensitive malignancies (pancreatic/ampullary) among all malignant cases exhibited a sharp and sustained increase, while the proportion of EUS-insensitive malignancies continued its previous trend. The formal DID regression analysis confirmed this visual evidence, revealing a significant and positive interaction term. The results indicated that the widespread adoption of EUS was associated with a substantial increase in the odds of diagnosing an EUS-sensitive malignancy compared to the EUS-insensitive group (details provided in subsequent analysis).

**Figure. 5.**
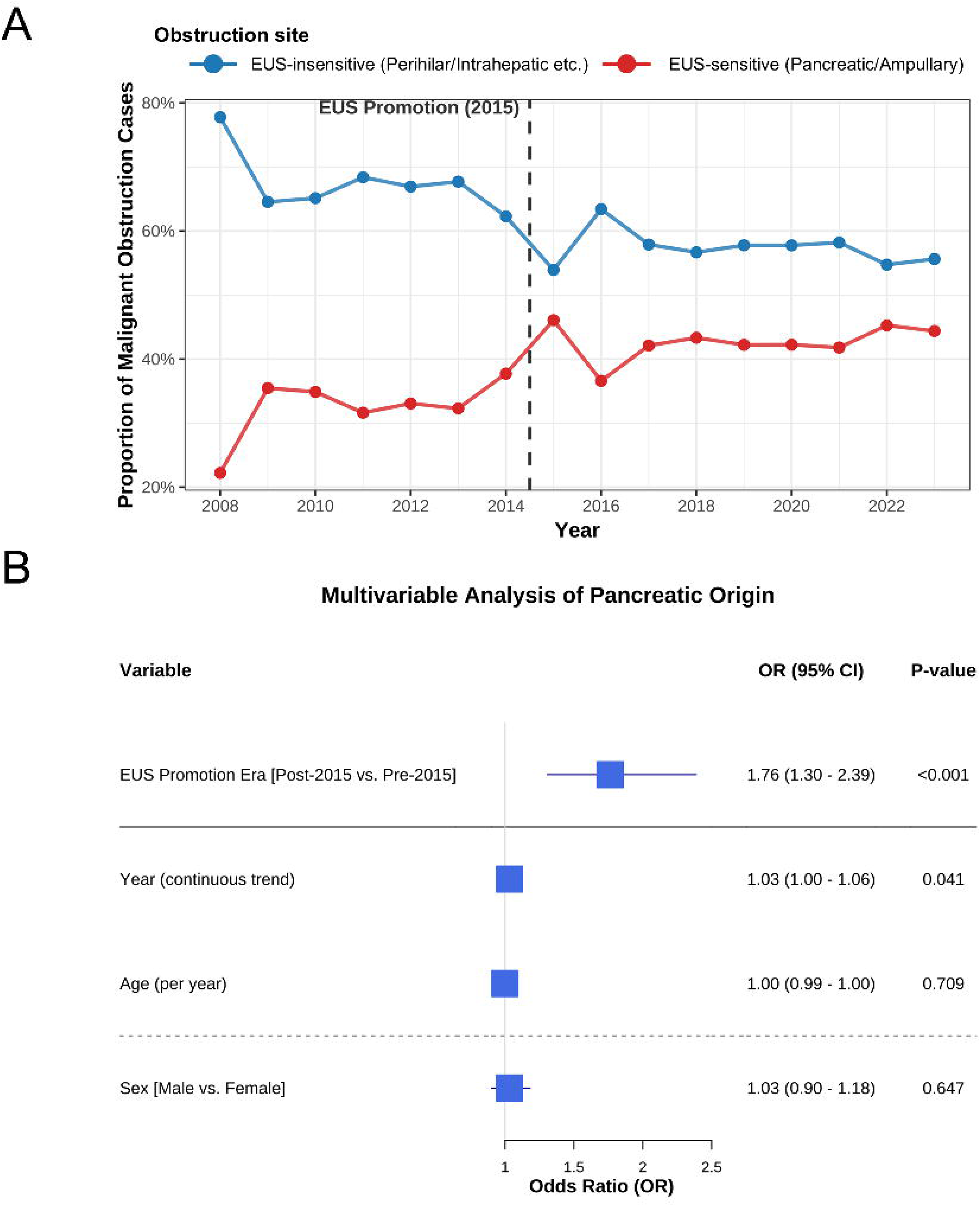
Difference-in-Differences (DID) analysis of the impact of EUS promotion on the diagnostic proportion of malignant biliary obstruction. **(A)** Trends in the proportion of malignant obstruction diagnoses for the EUS-sensitive group (pancreatic/ampullary obstruction) and the EUS-insensitive group (perihilar/intrahepatic obstruction). The dashed vertical line at 2015 marks the intervention point of EUS promotion. **(B)** Forest plot of the multivariable regression analysis for obstructions of pancreatic origin. The plot displays the odds ratios (ORs) and 95% confidence intervals (CIs) for factors associated with obtaining a malignant diagnosis, including the EUS promotion era (post-2015 vs. pre-2015), continuous year trend, age, and sex.

#### Factors associated with pancreatic-origin malignancy

To further substantiate the hypothesis that evolving diagnostic capabilities, rather than a true shift in disease incidence, drove the observed changes, we conducted a multivariable logistic regression analysis to identify factors associated with a malignant diagnosis of pancreatic origin (Figure 5B).

The analysis revealed that the EUS promotion era was a powerful independent predictor. Patients diagnosed in the post-2015 period had 1.76 times the odds of having a pancreatic-origin malignancy compared to those in the pre-2015 period (OR = 1.76, 95% CI: 1.30 - 2.39, p < 0.001). Furthermore, even after accounting for this era effect, the calendar year itself remained a significant predictor, with each passing year associated with a 3% increase in the odds of such a diagnosis (OR = 1.03, 95% CI: 1.00 - 1.06, p = 0.041). In contrast, patient age and sex were not significant predictors. These findings collectively suggest that the observed rise in pancreatic malignancies is strongly linked to the continuous evolution and adoption of advanced diagnostic technologies like EUS over the entire study period.

## Discussion

In this large, 15-year, single-center study, we document a notable shift in the apparent etiological spectrum of pathologically confirmed biliary obstruction, characterized by a steadily rising proportion of malignancy, particularly pancreatic cancer. A key finding, supported by a quasi-experimental Difference-in-Differences analysis, is that this trend does not appear to be an isolated epidemiological event. Instead, it appears to be closely associated with, and is plausibly driven by, a concurrent evolution in diagnostic technology which includes the ascendancy of Endoscopic Ultrasound (EUS). Our data therefore strongly suggest that the observed increase in specific malignancies may be substantially attributable to our improved ability to detect them; in essence, we have learned to see the disease better.

A plausible mechanism for this shift in diagnostic visibility is rooted in the unique capabilities of EUS. By placing a high-frequency ultrasound transducer in close proximity to the pancreas and distal bile duct, EUS provides superior spatial resolution compared to conventional imaging^[13]^. This allows for the detection and, crucially, the safe and accurate sampling of small lesions that might have been radiologically occult or inaccessible to other modalities like ERCP-guided brushing or percutaneous biopsy^[14]^. This technical superiority offers a compelling explanation for the specific increase in pancreatic cancer diagnoses observed in our cohort. It is highly plausible that a portion of these cases might have been previously misclassified as idiopathic obstruction, attributed to benign strictures, or simply remained undiagnosed in the pre-EUS era. Our findings thus call into question a simplistic interpretation of registry data that might point to a true and rapid surge in pancreatic cancer incidence as the consequence of this trend. While a genuine increase in disease incidence due to evolving risk factors cannot be entirely ruled out, our study provides strong evidence suggesting that a significant portion of the observed increase could be a diagnostic artifact, a phenomenon known as detection bias. This has important implications for the interpretation of historical and contemporary cancer epidemiology data, especially in fields undergoing rapid technological advancement. It serves as a reminder that observed trends in disease prevalence should always be interpreted within the context of concurrent shifts in diagnostic capabilities.

From a clinical and public health standpoint, the implications of our study extend beyond simply endorsing a technology. Clinically, our findings highlight the transformative impact of EUS-guided tissue acquisition on the diagnostic certainty of biliary obstruction. The crucial advantage demonstrated is not merely superior visualization, but the ability to secure a definitive histopathological diagnosis from lesions previously deemed inaccessible or too risky to sample. This capability is paramount, as it allows clinicians to move beyond presumptive diagnoses based on inconclusive imaging and low-yield cytology from ERCP. By reliably differentiating malignant strictures from benign mimics, EUS-guided biopsy directly facilitates the timely initiation of appropriate treatment, either neoadjuvant therapy for a confirmed pancreatic cancer or avoiding unnecessary surgery for an inflammatory condition. Our data therefore advocate for positioning EUS with biopsy capability not as a mere troubleshooting tool, but as a foundational step for establishing a precise etiological diagnosis whenever initial cross-sectional imaging is equivocal.

For health systems and hospital administration, these findings reframe the value proposition of advanced endoscopy. The diagnostic shift we observed suggests that an increasing caseload of diseases like pancreatic cancer is, in part, a direct return on investment in diagnostic precision. This distinction is critical: it implies that resource planning should not only react to an apparent rise in disease incidence but also proactively account for the increased diagnostic yield that advanced technologies provide. Accurately mapping the true, pathology-confirmed disease burden allows for more precise forecasting of the demand for highly specialized services, such as pancreatic surgery, interventional radiology, and oncology. Therefore, investing in EUS infrastructure and expertise is fundamentally an investment in data quality, providing a more accurate evidence base for strategic planning, quality improvement initiatives, and equitable resource allocation.

The major strengths of our study include its large sample size, long study duration spanning 16 years, and the robust focus on a pathology-confirmed cohort, which ensures the accuracy of our outcome measures. Furthermore, the application of a DID model provides a strong methodological framework for inferring causality from observational data.

However, certain limitations must be acknowledged. First, as a single-center study, our findings regarding the specific timeline and adoption rate of EUS may not be fully generalizable to other institutions with different resource levels or practice patterns, although the underlying principle is likely universal. Second, the retrospective design is inherently susceptible to information bias, though we sought to minimize this by using a structured electronic database. Finally, the strict exclusion of patients without a pathological report confirming the etiology inevitably introduces selection bias. By focusing only on pathologically confirmed cases, our cohort may not fully represent the real-world clinical heterogeneity of biliary obstruction and likely underestimates the proportion of rare or benign conditions that frequently lack pathological diagnosis, such as IgG4-related sclerosing cholangitis, post-lithiasis inflammatory strictures, parasitic infections, and HIV-cholangiopathy.

## Conclusion

In conclusion, the changing face of biliary obstruction over the past 15 years appears to reflect a complex interplay between potential epidemiological shifts and our evolving ability to characterize the disease with greater precision. Diagnostic progress, spearheaded by EUS-guided tissue acquisition, has likely facilitated the identification of a disease burden that was previously less visible, particularly in pancreatic cancer. While these observations offer only a limited perspective, they highlight that diagnostic evolution should be considered a meaningful factor influencing our understanding and management of this common clinical syndrome.

## Data Availability

All data produced in the present study are available upon reasonable request to the authors.

## Abbreviations

EUS: endoscopic ultrasound
EUS-FNA/B: Endoscopic Ultrasound-guided Fine-Needle Aspiration and/or Biopsy
DID: Difference-in-Differences
ERCP: endoscopic retrograde cholangiopancreatography
MRI/MRCP: magnetic resonance imaging/magnetic resonance cholangiopancreatography
CECT: contrast-enhanced computed tomography

## Acknowledgements

This work was financially supported by the National Natural Science Foundation of China (No. 82403592), the China Postdoctoral Science Foundation (No. 2024M762234), and the Sichuan Science and Technology Program (No. 2024NSFSC1899).

## Declaration of interests

Authors disclose no conflict of interest for this study.

## Author contributions

Study concept and design: Ningyuan Wen and Yonggang Wei; Data acquisition: Ningyuan Wen, Nianheng Wu and Haojun Wu; Data analysis and interpretation: Ningyuan Wen and Haili Zhang; Drafting of the manuscript: Ningyuan Wen, Yufu Peng and Hongwei Xu; Funding: Yonggang Wei. All authors have read and critically revised the manuscript and agreed to the published version.

---

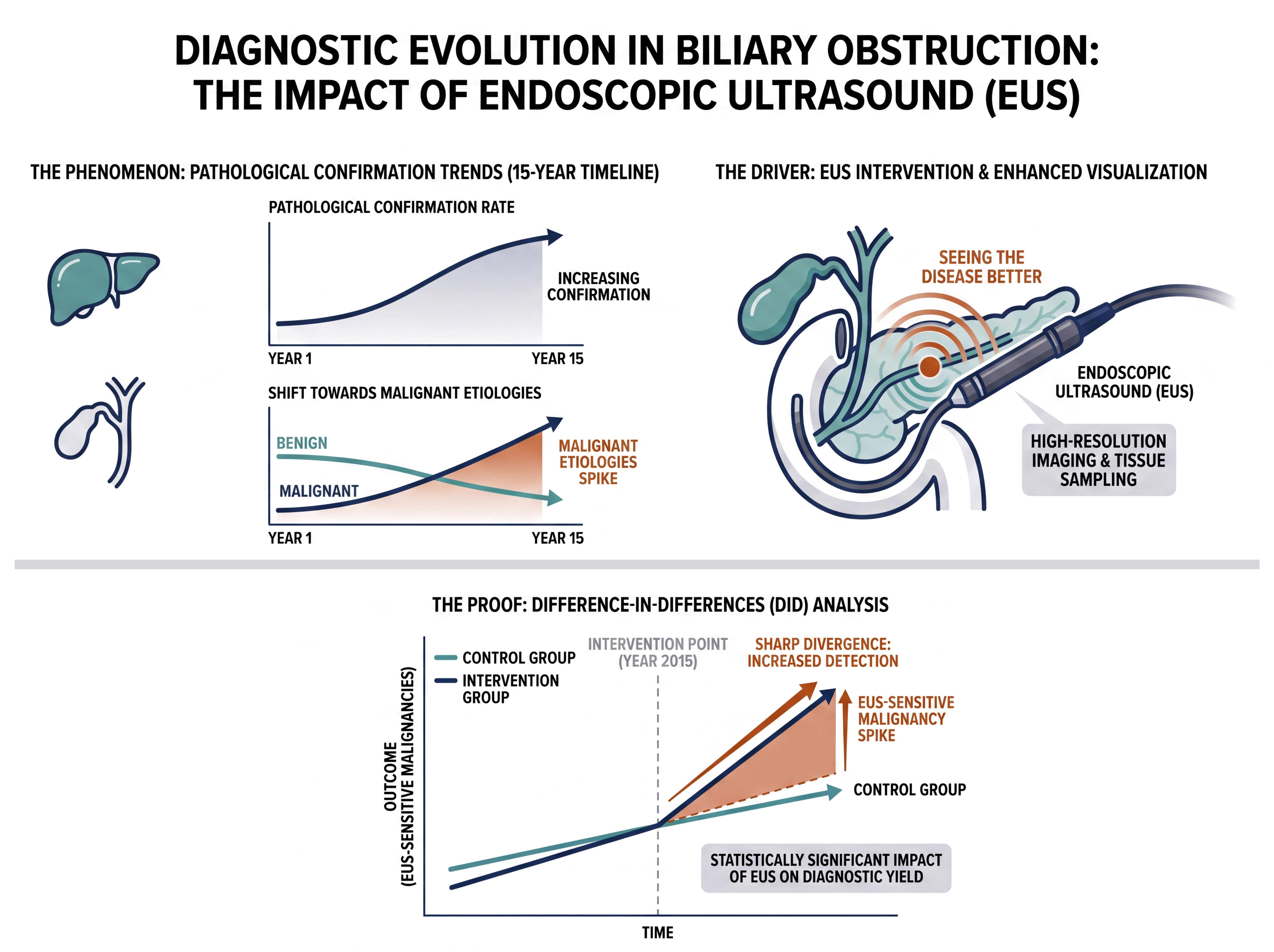

## Notes

### Competing Interest Statement

The authors have declared no competing interest.

### Author Declarations

The study protocol was approved by the Ethics Committee on Biomedical Research, West China Hospital of Sichuan University, and the requirement for individual informed consent was waived due to the retrospective nature of the analysis. The study was preregistered in the Open Science Framework (registration DOI: https://doi.org/10.17605/OSF.IO/JYDS3) to ensure transparency.

