## Supplementary material for "The Impact of Endoscopic Ultrasound Adoption on Etiological Shifts in Biliary Obstruction: A 15-Year Real-World Study": Table 1

Table1. Baseline demographic and clinical characteristics of the study cohort, stratified by study period.

| **Characteristic** | **Overall**  N = 5672 (100%)^1^ | **2008-2015**  N = 1262 (22%)^1^ | **2016-2023**  N = 4410 (78%)^1^ | **p-value**^2^ |
| --- | --- | --- | --- | --- |
| **Age (years), Median (Q1 - Q3)** | 58.4 (49.0 - 67.2) | 58.2 (48.1 - 66.8) | 58.5 (49.2 - 67.3) | 0.24 |
| **Sex, n (%)** |  |  |  | 0.88 |
| Female | 2,448 (43.2%) | 547 (43.3%) | 1,901 (43.1%) |  |
| Male | 3,224 (56.8%) | 715 (56.7%) | 2,509 (56.9%) |  |
| **Pathology Type, n (%)** |  |  |  | **<0.001** |
| Benign | 1,978 (34.9%) | 517 (41.0%) | 1,461 (33.1%) |  |
| Borderline | 11 (0.2%) | 5 (0.4%) | 6 (0.1%) |  |
| Malignant | 3,683 (64.9%) | 740 (58.6%) | 2,943 (66.7%) |  |
| **Main Etiologies, n (%)** |  |  |  | **<0.001** |
| Common bile duct stones | 1,039 (18%) | 315 (25%) | 724 (16%) |  |
| Pancreatic ductal adenocarcinoma | 973 (17%) | 188 (15%) | 785 (18%) |  |
| Hilar cholangiocarcinoma | 565 (10.0%) | 90 (7.1%) | 475 (11%) |  |
| Distal cholangiocarcinoma | 595 (10%) | 135 (11%) | 460 (10%) |  |
| Acute pancreatitis | 327 (5.8%) | 58 (4.6%) | 269 (6.1%) |  |
| Other diseases | 2,173 (38%) | 476 (38%) | 1,697 (38%) |  |
| **EUS-guided FNA/FNB Used, n (%)** | 859 (15%) | 139 (11%) | 720 (16%) | **<0.001** |
| **Total Bilirubin (μmol/L), Median (Q1 - Q3)** | 66.8 (46.5 - 110.6) | 64.5 (52.4 - 122.3) | 67.7 (42.7 - 109.2) | **<0.001** |
| **Direct Bilirubin (μmol/L), Median (Q1 - Q3)** | 39.6 (22.1 - 72.9) | 27.5 (18.7 - 81.9) | 41.4 (24.4 - 71.9) | **<0.001** |
| **CA 19-9 (U/mL), Median (Q1 - Q3)** | 71.8 (23.2 - 250.0) | 45.3 (17.8 - 175.5) | 82.6 (25.1 - 269.3) | **<0.001** |
| ^1^Data are presented as median (IQR) or n (%). | | | | |
| ^2^P-values calculated using Wilcoxon rank-sum test for continuous variables and Chi-squared test for categorical variables. | | | | |
